# A Local Outpatient Practice-Level Prediction Model for Short-Term Psychiatric Emergency Presentation

**DOI:** 10.64898/2026.06.29.26356785

**Authors:** John L. Havlik, Ben Tyrrell, Jeanette Polaschek, Nathan Bell, Eric R. Arzubi

**Affiliations:** Department of Psychiatry and Behavioral Sciences, Stanford University School of Medicine, Palo Alto, CA; Big Sky Care Connect, Helena, MT; Frontier Psychiatry, Billings, MT; Yale Child Study Center, Yale University School of Medicine, New Haven, CT

## Abstract

**Importance:** Psychiatric emergency department (ED) presentations are difficult to predict using general medical risk stratification tools. Health information exchange (HIE) data may improve prediction by capturing fragmented care across settings.

**Objective:** To develop and temporally validate a machine learning model using HIE and geospatial data to predict 30-day psychiatric ED presentation among outpatients receiving psychiatric care and to compare its performance with standard clinical risk scores.

**Design, Setting, and Participants:** This retrospective cohort study included patients seen at Frontier Psychiatry with records in the Big Sky Care Connect statewide HIE. Structured clinical data were linked to zip code–level sociodemographic measures. The analytic unit was the patient snapshot, defined as all structured data available up to a given point. Models were evaluated in temporally separated train and test sets.

**Exposures:** Predictors derived from HIE structured data, including prior utilization, diagnoses, medications, laboratory data, and zip code–linked geospatial deprivation and vulnerability measures.

**Main Outcomes and Measures:** The primary outcome was psychiatric ED presentation within 30 days, identified from structured encounter-type fields and primary diagnosis codes for psychiatric or substance use disorders. Model discrimination was compared with a parsimonious clinical baseline model and LACE and Elixhauser scores.

**Results:** In the test set, 343 of 16,469 snapshots (2.1%) were followed by a qualifying psychiatric ED presentation within 30 days, corresponding to 102 ED visits among 68 patients. The machine learning model showed discrimination in temporally held-out testing and outperformed the clinical baseline model as well as LACE and Elixhauser scores. At a prespecified decision threshold, the model reduced the number needed to evaluate from more than 40 with universal screening to 3.4 to identify 1 true-positive case, while identifying over two fifths of 30-day psychiatric ED presentations.

**Conclusions and Relevance:** In this retrospective cohort study, a locally developed machine learning model using statewide HIE data showed improved prediction of 30-day psychiatric ED presentation compared with selected general-purpose risk scores. The results support the feasibility of HIE-enabled local psychiatric risk modeling and suggest other practices could develop similarly tailored models. Prospective studies are needed to assess clinical utility and effects on outcomes.

## INTRODUCTION

Predicting psychiatric emergencies is a large and unsolved problem. As of now, care structures are largely focused on acute, expensive, and reactive care.^1^ Psychiatric emergency department (ED) visits are a trying and often very expensive experience for patients and payors. It is possible that many of these ED visits reflect issues which could have been treated in an outpatient setting, but have spiraled to the point that serious harm may occur and emergency psychiatric evaluation and potential inpatient stabilization is warranted. Psychiatric patients often have a difficult time advocating for appropriate care in the outpatient setting because navigating the fragmented, “ghost provider” rich^2^ and narrow^3,4^ mental healthcare network landscape is challenging. Predicting psychiatric emergency needs to enable proactive response would thus be a welcome solution to a pressing societal problem.

There are limited attempts in the literature to solve the problem of predicting psychiatric emergencies. Some recent adjacent literature has attempted to predict mental health symptom risk after general inpatient hospitalization.^5^ While there are more general estimates of comorbidity and readmissions risks such as the Elixhauser score^6^ or LACE index^7^ that are commonly used to risk stratify patients, psychiatric-specific risk stratification remains unaddressed. Risk stratifying to figure out when to proactively intervene before psychiatric emergencies occur would mark a major shift in the field’s abilities to provide timely and value-based care to patients who need it most, before harm occurs.

Recent necessary advances on both the technical and healthcare infrastructure fronts may have enabled the creation of such a tool. In machine learning, advances include interpretable and highly effective supervised learning techniques such as logistic regression, random forest modeling, and gradient boosting. Such machine learning techniques may enable meaningful insights into patient risk even when trained using only patients receiving care from a single specialty practice. In healthcare infrastructure, governments have begun to facilitate interoperability of electronic healthcare records data across a variety of healthcare settings through the creation of healthcare information exchanges (HIEs). In Montana, Big Sky Care Connect (BSCC) was founded in 2018 as a 501(c)(3) nonprofit to respond to the need for a statewide coordinated health information exchange. BSCC is the state designated HIE in Montana and shares healthcare data across hospitals, clinics, FQHCs, pharmacies, and other healthcare settings throughout the state and works closely with the Montana Department of Health and Human Services.^8^ BSCC is an opt-out provider of healthcare information with 3,120 unique providers (including physicians, nurses, and other clinical staff) and 1,702,991 unique patients served as of May 2026.

The present paper describes the development and evaluation of a supervised machine learning model to predict 30-day psychiatric ED visits. This model was trained on HIE data from patients receiving care from a psychiatry practice in Montana (Frontier Psychiatry). Because psychiatric emergency risk is shaped by local care networks and service availability, we designed this study as a proof of concept for local prediction modeling rather than as development of a universally transportable model. Accordingly, our objective was to assess whether routinely available local HIE data could support short-term risk stratification within this specific care setting. We emulated a prospective trial of our model by training on all practice patients entering the HIE before a certain date, and testing on all patients entering the HIE after this date. We hypothesized that our model would outperform general-purpose readmission and comorbidity indices on this task. We further sought to determine whether any predictive advantage from higher-dimensional machine learning models would justify their added complexity relative to a parsimonious clinical model.

## METHODS

### Institutional approval and participant consent

This study received Institutional Review Board approval from the WIRB-Copernicus Group Inc. institutional review board. Informed consent was not required as data were retrospective and de-identified according to standard privacy guidelines; no patients or the public were involved in study design. This study followed the TRIPOD+AI reporting guideline for the development and validation of prediction models in healthcare.^9^

### Data sources and study population

To develop predictive models for 30-day emergent psychiatric presentations, we constructed a retrospective cohort using multi-source clinical data of all patients seen by Frontier Psychiatry with records in the Big Sky Care Connect (BSCC) healthcare information exchange (HIE) database. This dataset contains datapoints such as patient encounters, labs, vitals, and other data. A full data dictionary for the variables derived from the BSCC used in this study can be found in **Supplemental Table 1**. Cognizant of place-based disparities in psychiatric access, we supplemented this HIE data with two established geospatial datasets, the Social Vulnerability Index (SVI)^10^ and Area Deprivation Index (ADI)^11^, linking patients to SVI and ADI values based on zip code values from the HIE. Each observation consisted of a patient “snapshot,” defined as all structured data available up to a given index date. Final sample size, number of snapshots, and event counts are reported in Results.

### Anchor Date, Index Date, and Snapshot Definitions

We defined each patient’s anchor date in the HIE as the earliest recorded interaction across all available structured data sources, including encounters, laboratory results, medication prescriptions, vital signs, and diagnoses. Patients were then assigned to temporally distinct cohorts using a cutoff date corresponding to the 90th percentile of anchor dates: a training cohort comprising patients with any anchor dates before June 5, 2023, and a test cohort comprising patients with anchor dates on or after June 5, 2023. This patient-level temporal split, based on HIE data available through October 30, 2025, was chosen to approximate prospective deployment and reduce information leakage;^12,13^ accordingly, no patient contributed snapshots to both training and test sets. Outcome prevalence in the test cohort was left unchanged to provide realistic estimates of model performance.

We transformed each patient’s longitudinal record into multiple prediction snapshots, generating one snapshot for each encounter that met minimum observation criteria. Each snapshot was defined as a patient encounter on a given date, with all predictors derived exclusively from data recorded before that index date. All encounters were eligible to serve as index events. Details on snapshot exclusions can be found in our eMethods in **Supplement 1.**

### Primary Outcome Definition

The primary outcome was an emergency department (ED) presentation for a psychiatric or substance use–related condition within the subsequent 30 days. ED presentations were identified using encounter type fields in the HIE “Encounter” datafile indicating an encounter type of “Emergency”. We defined an encounter for psychiatric reasons as an encounter with a primary International Classification of Diseases, Tenth Revision (ICD-10) diagnosis codes outlined in the eMethods in **Supplement 1.**

### Secondary Benchmarking Definitions

Secondary benchmarks included the LACE index and Elixhauser comorbidity index, which were selected as established general-purpose measures of readmission risk and comorbidity burden. Because general-purpose indices such as LACE and Elixhauser are not designed for psychiatric emergency prediction, we also specified a parsimonious clinical baseline model using a small set of face-valid predictors available in routine practice. Predictors were age, sex, race, ethnicity, number of mental health diagnoses in the prior 90 days, any psychiatric ED visits in the prior 90 days, and total encounters in the prior 90 days. This logistic regression model was tuned, isotonic scaled, and evaluated within the same nested cross-validation and temporally held-out testing framework as the primary model.

### Feature engineering

We derived candidate predictors from structured data available before each snapshot index date using a domain-based feature engineering pipeline designed to prevent temporal leakage. Features were grouped into utilization, diagnostic, pharmacy, physiology/laboratory, demographic, and social determinant domains. Missing values were retained during feature construction and addressed subsequently within model-specific preprocessing. See the eMethods in **Supplement 1**.

### Model Comparator Specification

Before evaluation in the temporally held-out test set, we specified the primary outcome, candidate comparator models, and the model operating point. First, we developed a portfolio of machine learning models, including Logistic Regression, Random Forest and LightGBM, to capture linear, bagging, and boosting signal structures, respectively. To tune hyperparameters and conduct preliminary performance testing for all models, we utilized a 5x5 nested cross-validation (NCV) framework^14,15^ on the training set with limited hyperparameter grid search performed on the inner folds for all models. See the eMethods in **Supplement 1**.

### Model Testing

Our operating goal was to catch the top third of emergency psychiatric presentations while maximizing precision. This operating point target was informed by Frontier’s capacity for proactive outreach and consistent with recommendations that prediction thresholds be aligned with intended clinical use and intervention burden.^16^ We determined our testing decision threshold using the hyperparameter tuning described above, plus an additional temporal adjustment for trailing 180-day event prevalence. Once our final model was selected, we tested our hyperparameter-tuned classifier on our temporally-separated testing data; details of this testing are outlined in eMethods in **Supplement 1.**

To measure clinical utility, we performed Decision Curve Analysis (DCA) to quantify net benefit across a range of decision thresholds, with specifications outlined in our eMethods in Supplement 1. To provide an interpretable window into the risk of algorithmic bias, we assessed model performance (operationalized by area under the precision-recall curve) stratified by sex, race, ethnicity, and age band, with a prespecified subgroup sensitivity analysis accounting for differences in base rate prevalences. We additionally interrogated top model features using SHAP (SHapley Additive exPlanations) values to generate global beeswarm and local waterfall plots.

We conducted a limited set of statistical analyses in this manuscript, using DeLong’s test for correlated ROC curves^17^ to compare model performance between our final model, our parsimonious clinical baseline model, and LACE and Elixhauser scores (using their risk scores mapped to decisions across the same threshold grid), across all snapshots of patients with available risk score data. Significance was determined using a P<0.05, Bonferroni-Holm adjusted at the comparison group level.^18^ All confidence intervals reported in this study were generated via 1,000 bootstrap iterations with replacement, clustered at the patient level. Python version 3.14.3 was used for all analyses and for figure creation.

## RESULTS

### Study population

The analytic unit for this study was the patient snapshot. Our final study population consisted of 11,463 patients in our training set and 1,739 patients in our test set, comprising 143,203 training snapshots and 16,469 testing snapshots **Table 1, Supplemental Figure 1.** Our overall sample was 62.8% female, had a median [IQR] age of 36.6 [27.8-49.0], and had a median of 0 [0-0] emergency psychiatric presentations in the last year. In the test set, 343 snapshots (2.1%) were followed by a qualifying psychiatric ED presentation within 30 days, corresponding to 102 ED visits among 68 unique patients. Because patients could contribute multiple snapshots and multiple snapshots could precede the same event, snapshot-level event counts exceed unique event counts.

**Table 1.**
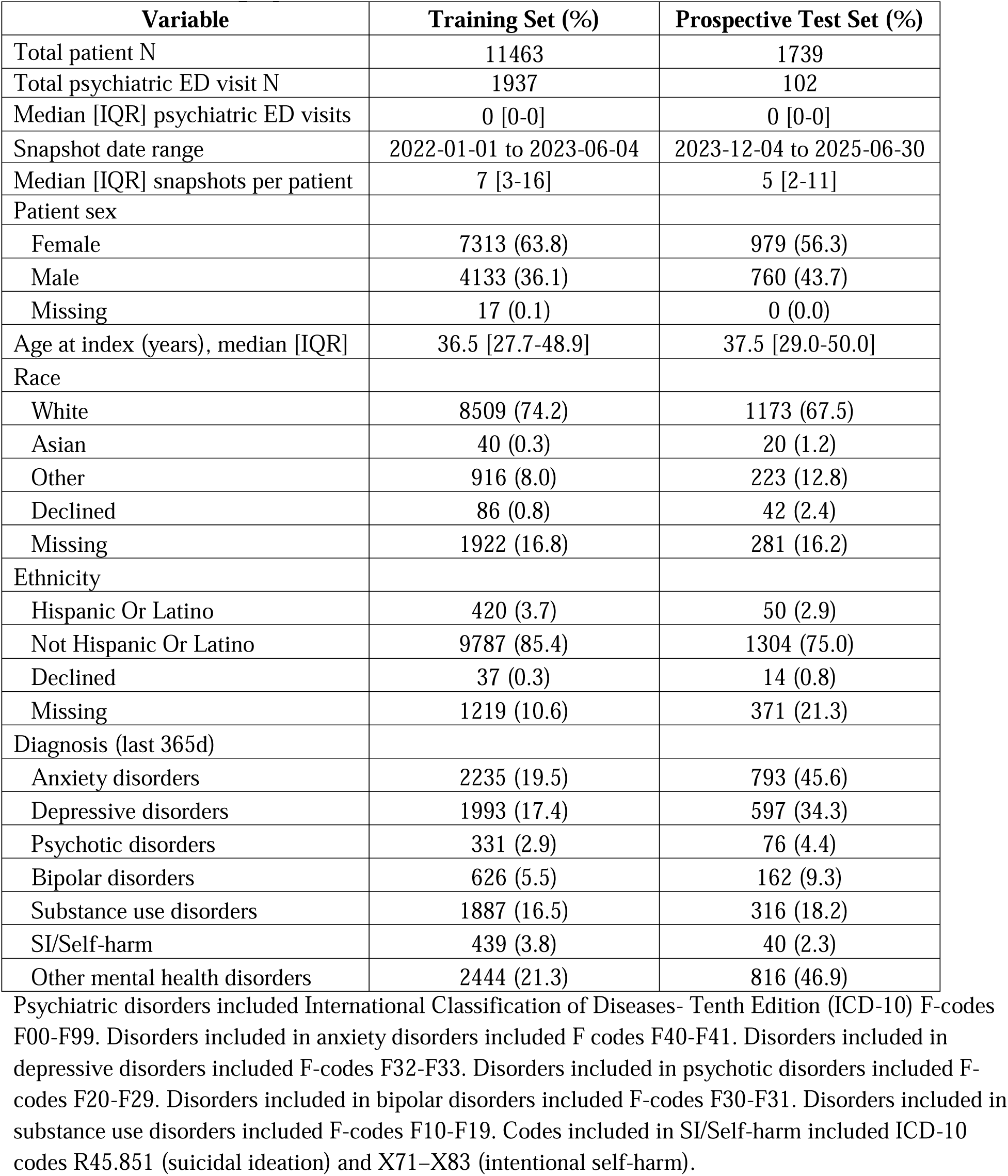
Patient Demographics and Event Information.

### Model selection

Our best-performing model on preliminary nested cross-validation was a random forest model, see **Supplemental Table 2** for preliminary cross-validation testing performance of all three models at optimized hyperparameters. This model had a mean ± standard deviation (SD) precision at 1/3 recall of 0.22 ± 0.05 on our fivefold outer-fold cross-validation testing, with a mean ± SD ROC AUC of 0.78 + 0.02 and PR AUC of 0.22 ± 0.07. The modal hyperparameter settings for this model across outer folds were a class weight of null, a max tree depth of 12, a maximum feature setting of the square root of all features, a minimum samples per leaf setting of 4, and an estimator number setting of 500. ***Performance on unseen test data***

In a prospective trial on all patients with anchor dates after the cutoff date, we applied the mean decision threshold which gave us 1/3 recall during the outer folds of our cross-validation testing. At this threshold, the model had a precision ± bootstrapped standard deviation (95% CI) of 0.29 (0.12-0.44) and identified 49.3% (21.1%-69.8%) of emergency psychiatric visits. This model would need to flag 3.4 (2.3-8.1) patients to identify one true positive emergency psychiatric visit within the next 30 days; full model metrics can be seen in **Supplemental Table 3**. Our prospectively trialed model had an ROC AUC of 0.85 (0.75-0.91) and PR AUC of 0.29 (0.08-0.48) on unseen test data. At this threshold, our model was 97.5% (96.6%-98.3%) specific and generated 3.5 (2.1-5.3) alarms per 100 patients (**Figure 1**). A sensitivity analysis using one random snapshot per test patient produced qualitatively similar performance metrics, **Supplemental Table 4.**

**Figure 1:**
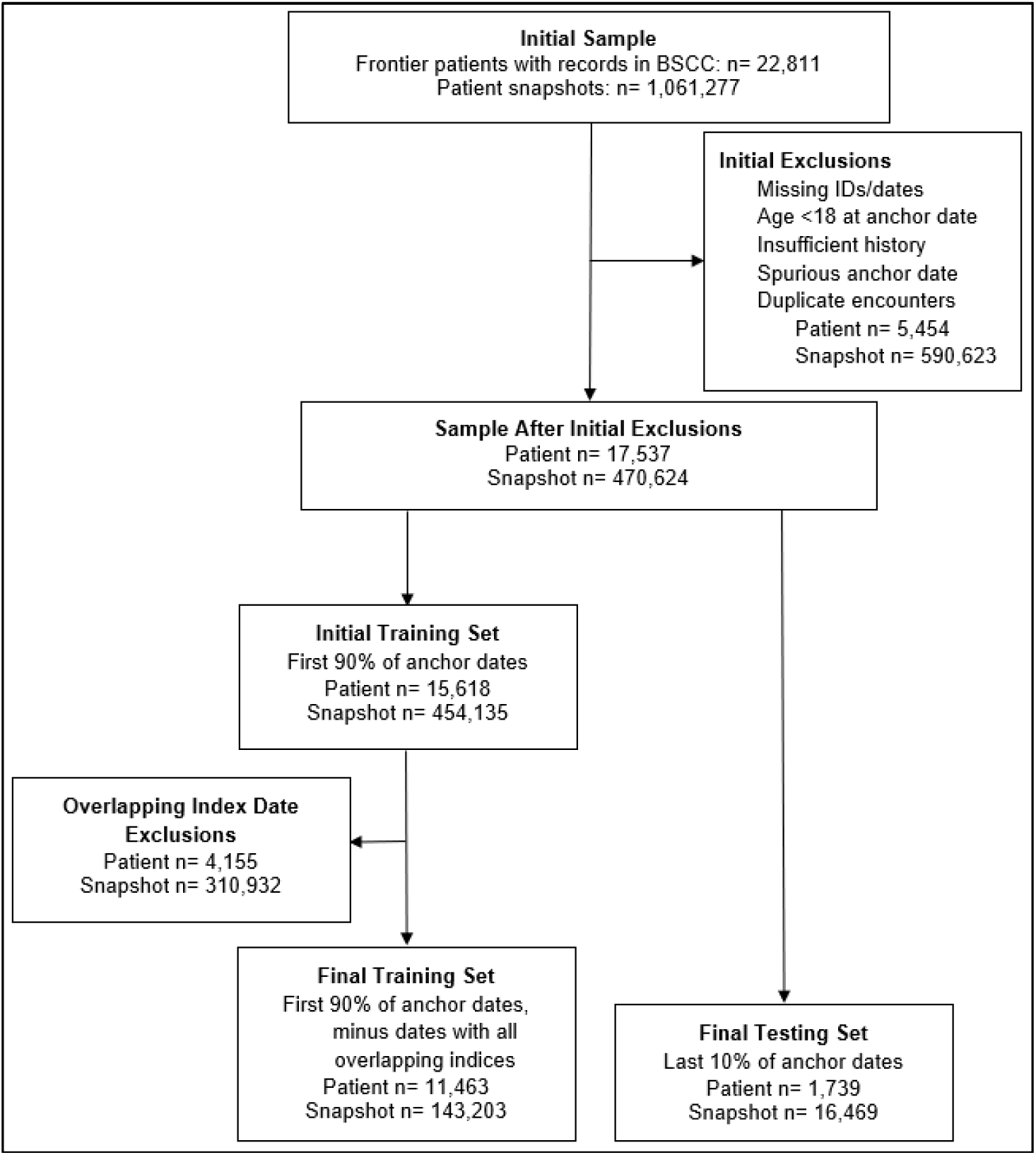
Study Sample and Exclusions. BSCC: Big Sky Care Connect. Patients were excluded if missing a unique patient identifier, if they were aged under 18 at time of their first patient record in BSCC (“anchor date”), if their anchor date predated BSCC creation; duplicate encounters and encounters with missing dates were also excluded. From this initial sample, we split our data at the patient level into patients with anchor dates in the first 90% chronologically (before 2023-06-05) and those with anchor dates after this date.

### Calibration analysis

Our calibration testing indicated our model’s ability to correctly gauge clinical risk was reasonable though imperfect on the test set (Brier=0.017; slope=1.36; intercept=1.08). In the low probability region (0–0.03), observed event rates tracked predicted probabilities, but global metrics indicated some overconfidence at higher probabilities (**Figure 2a**).

### Clinical utility: decision curve analysis

On decision curve analysis, our prespecified decision threshold implied a cost:benefit ratio of 0.09, meaning the cost of a false positive alarm was valued at approximately 11 times less than the benefit of a true positive alarm. At this operating point, the model’s net benefit was 8.0 per 1000 patients (95% CI: 1.0-19.4) relative to a baseline of no detection system. Decision curves were similar whether or not we *a priori* recalibrated our decision threshold for trailing 180-day event prevalence (**Figure 2b**).

**Figure 2:**
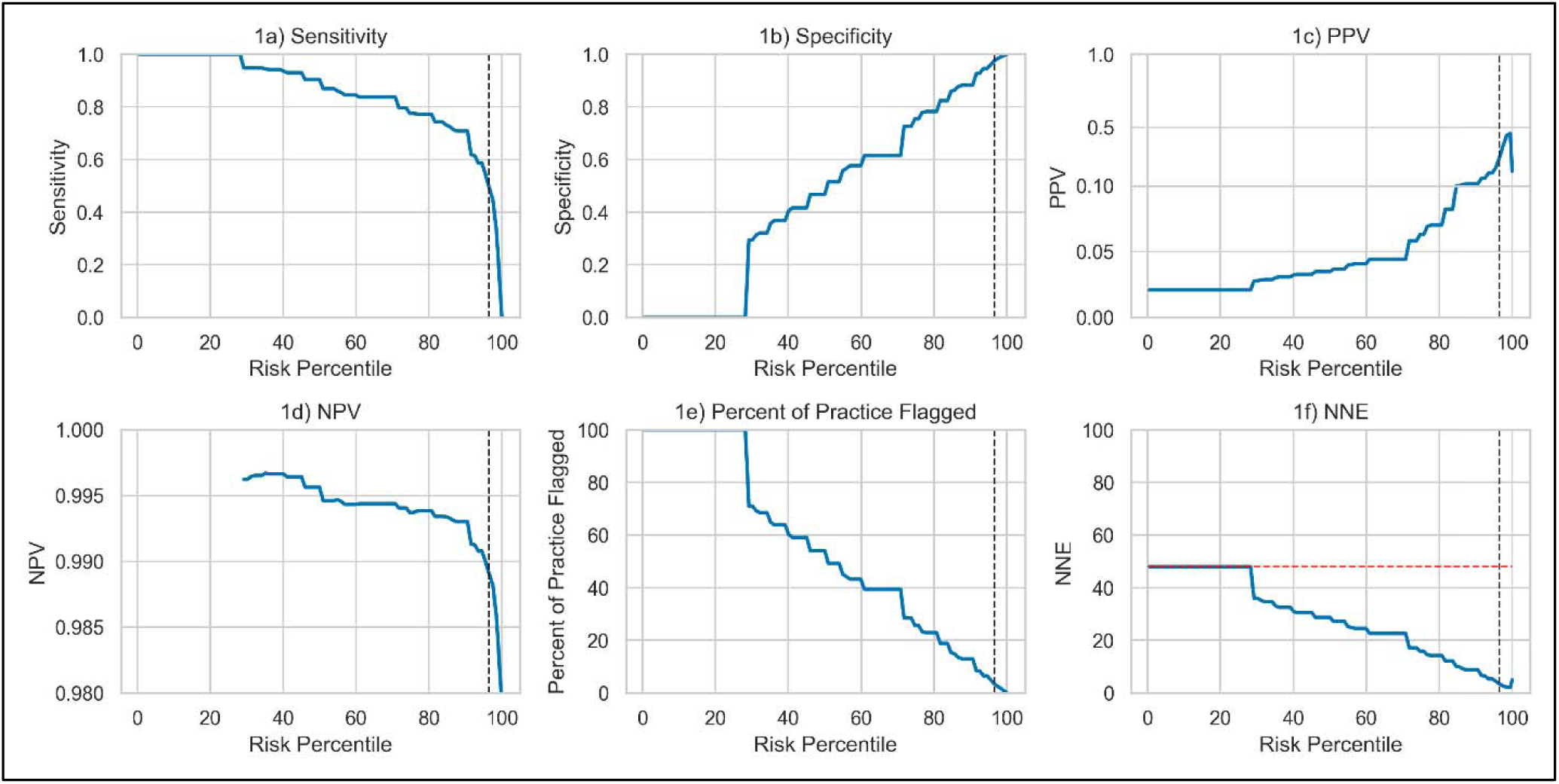
Model metrics across calculated risk percentiles. PPV: positive predictive value; NPV: negative predictive value; NNE: number needed to evaluate. A: Sensitivity on the held out test set as a function of risk percentile (higher percentiles correspond to more stringent thresholds). Vertical lines throughout this and subsequent figures mark sensitivity targets at prespecified decision threshold (dashed) targeting 1/3 recall. B: Specificity vs risk percentile. C: PPV vs risk percentile shown on split y axes to reveal detail at low values (lower pane: 0–0.1; upper pane: 0.1–1.0). D: NPV vs risk percentile, with a red dashed no skill reference equal to 1 − prevalence. E: Percent of practice flagged (alarms per 100 patients) vs risk percentile. F: NNE vs risk percentile, with a red dashed no skill reference equal to 1/prevalence; lower values indicate fewer false positives per true positive.

**Figure 2:**
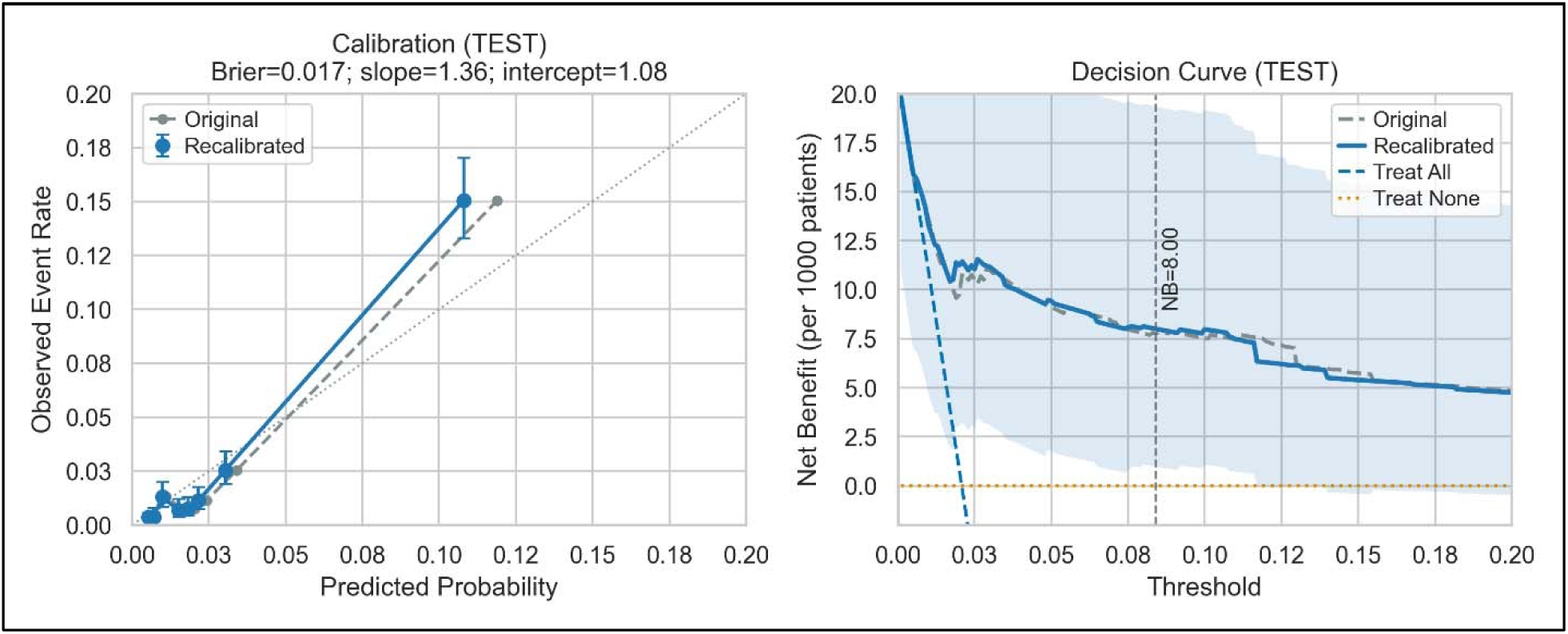
Model decision and calibration curves. NB: net benefit. A: Decision curve analysis on the test set shows net benefit per 1000 patients as a function of threshold in the low-risk range. Vertical line indicates operating point at prespecified decision threshold; this decision threshold implied a cost:benefit ratio of 0.09, meaning the cost of a false positive alarm was valued at approximately 11 times less than the benefit of a true positive alarm. Net benefit is calculated as true positives per patient minus preference weighted false positives per patient, scaled to 1000 patients; higher values indicate greater clinical utility. Light blue shading reflects the bootstrapped 95% confidence interval (CI) on the recalibrated test curve; the original (train prior) curve is shown as a dashed reference. B: Calibration panel which shows observed event rates vs predicted probabilities with 95% Wilson CIs; the recalibrated model tracks the 45° line in the low probability region.

### Subgroup analysis and model interpretability

Overall model performance was within 95% CIs for PR-AUC for all subgroups except male sex and those ages 18-24 (the youngest age group in our sample) and 65+ (the oldest age group in our sample), **Figure 3**. In these groups, the model’s discriminatory ability was worse than average across the entire cohort. These differences in discriminatory ability were robust to subgroup analyses using prevalence adjusted average precision in all cases (**Supplemental Figure 2)**.

Alongside this modeling, we conducted SHapley Additive exPlanations (SHAP) analysis to determine feature importance. Our models revealed top contributing features outlined in **Figure 4**. Notably, measures of utilization made up 14 of the 20 most important features in our model.

**Figure 3:**
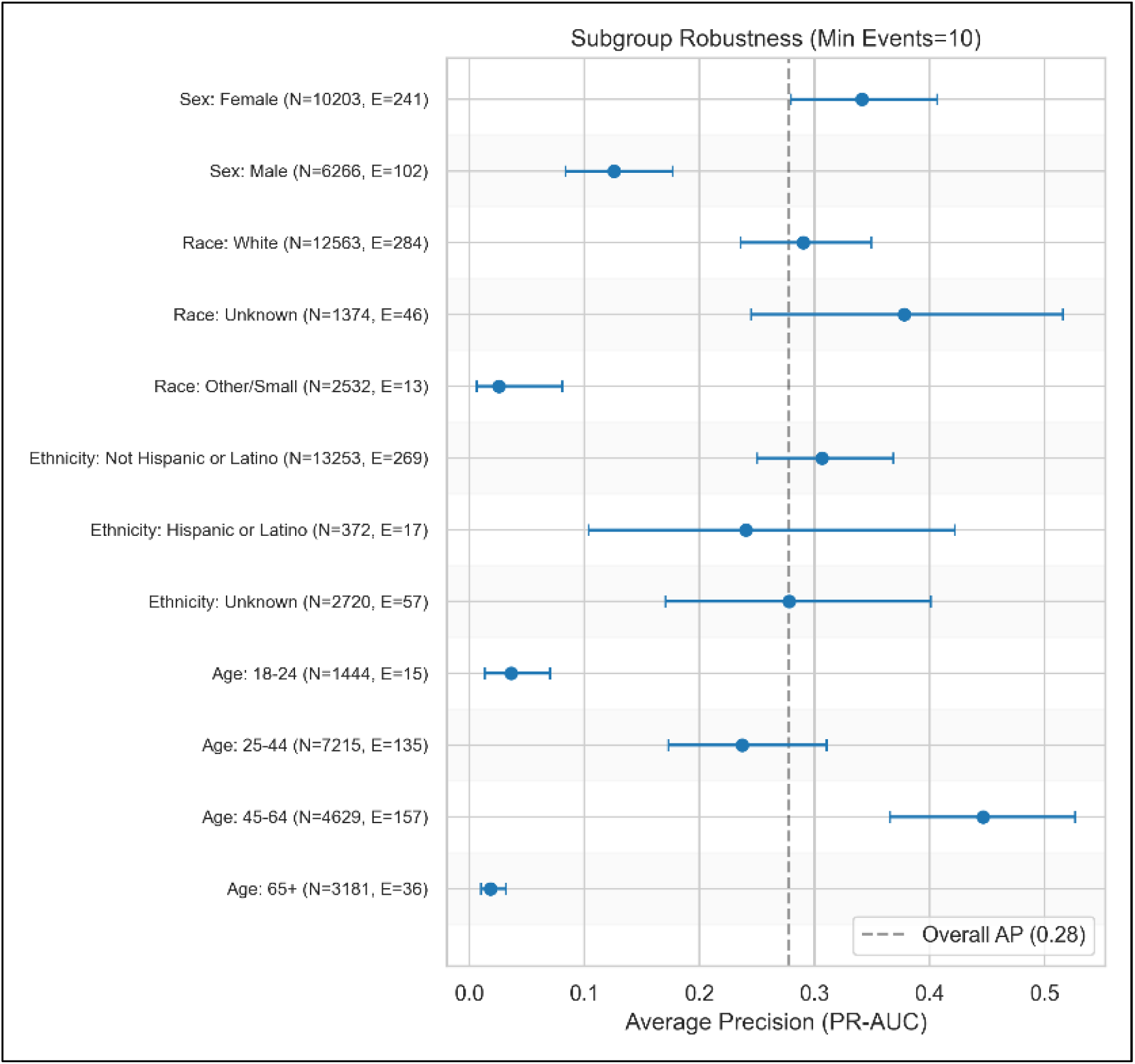
Model subgroup robustness analysis. Subgroup analysis conducted on age, sex, and race at the snapshot level; event denotes a snapshot with a 30-day emergency psychiatric visit. Points show subgroup performance with bootstrapped 95% confidence intervals. Dashed line denotes overall sAP. Higher values indicate better performance; sAP=0 corresponds to random ranking. Prevalence-adjusted estimates of subgroup performance can be seen in **Supplemental Figure 2.**

**Figure 4:**
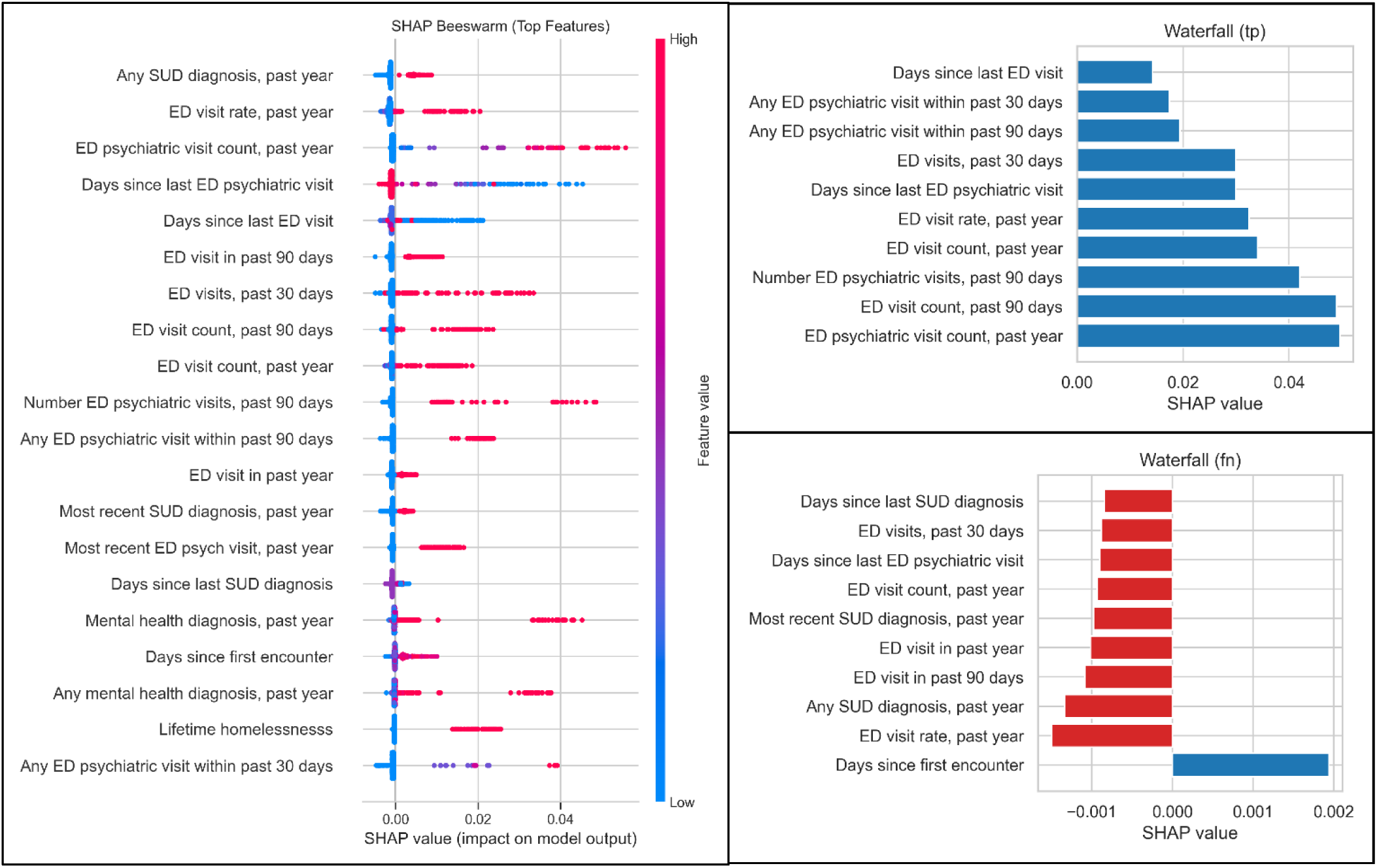
Model SHAP analysis. SHAP: SHapley Additive exPlanations; TP: true positive; FN: false negative; SUD: substance use disorder, ED: emergency department. A: SHAP beeswarm on the held out test set shows the distribution of feature contributions for the top predictors; features are ordered by mean absolute SHAP, dot color encodes feature value, and points to the right indicate contributions toward higher predicted risk. B: Waterfall plot for a representative high risk TP and for C: a missed FN highlight the top contributing features and their direction of effect for each case; bars to the right increase predicted risk and bars to the left decrease predicted risk, illustrating features behind individual predictions.

### Performance versus generalized risk scoring models

The final model’s ROC AUC was higher than the parsimonious clinical model as well as LACE and Elixhauser score stratification: vs clinical baseline ΔAUC=0.10 (95% CI 0.07–0.13; P<0.001); vs LACE ΔAUC=0.19 (95% CI 0.08–0.28; P<0.001); vs Elixhauser ΔAUC=0.27 (95% CI 0.14–0.37; P<0.001), **Figure 5**. Interrogating differences in risk scoring distribution versus Elixhauser and LACE risk scoring of our test cohort, both our final model and the parsimonious clinical baseline showed a conservative and right-skewed approach to emergent psychiatric presentation probability assessment, versus a qualitatively more normalized distribution for Elixhauser and LACE scoring, **Figure 6a**. The final model was effective in separating probabilities of those with emergent psychiatric presentations from those without emergent psychiatric presentations, (ΔProbability=0.11 (95% CI 0.04–0.17; P<0.001, **Figure 6b**).

**Figure 5:**
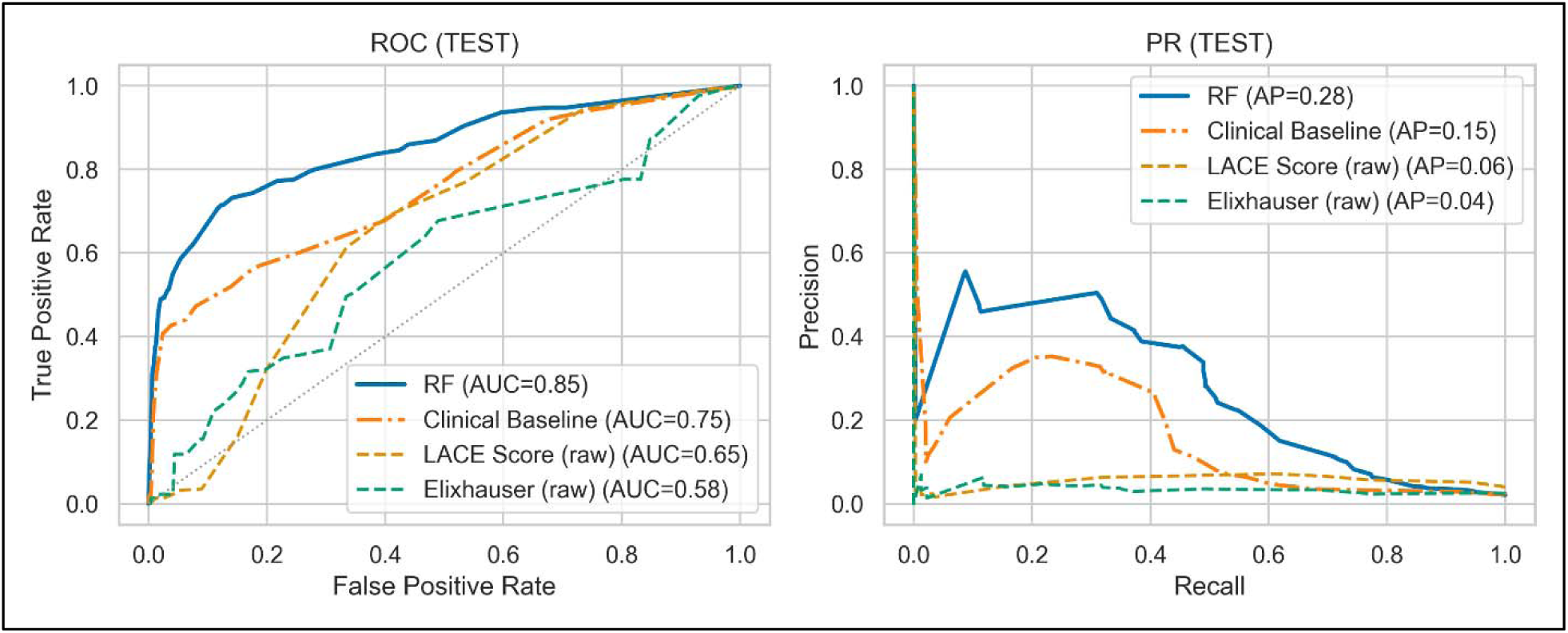
Model ROC and PR cuves. AUC: area under curve; ROC receiver-operator characteristic; PR: precision-recall; RF: random forest; AP: average precision. A: ROC curve, optimal model versus Elixhauser and LACE scores. B: PR curve, optimal model versus Elixhauser and LACE scores.

**Figure 6:**
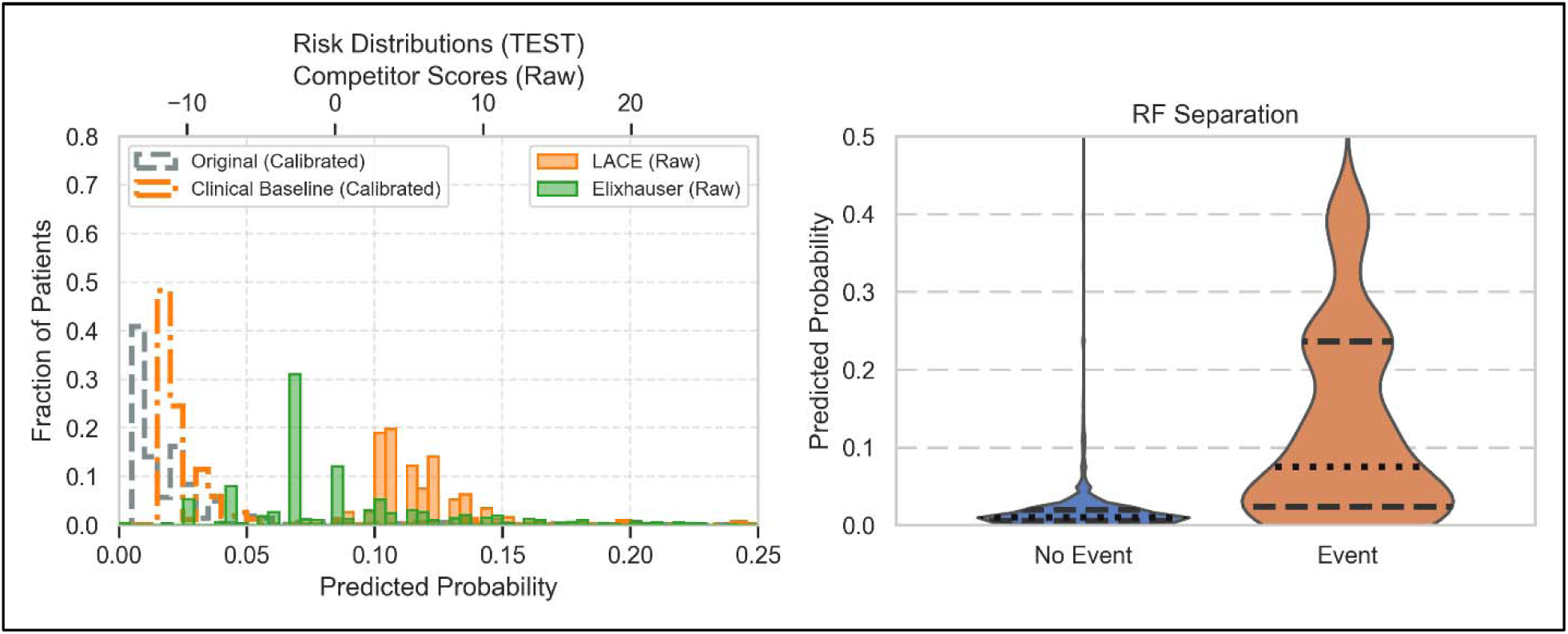
Model risk distributions versus generalized models (A) and risk distribution by event status (B). RF: random forest. A: scoring distribution comparison, optimal model versus LACE and Elixhauser scores. Bottom X axis shows predicted probability of emergent psychiatric presentation from optimal model. Top X axis shows scoring of Elixhauser and LACE score comparators. B: Violin plot of optimal model probability distributions. Dashed lines represent median probabilities; dotted lines represent interquartile range probabilities. Probabilities were lower (mean 0.02, SD 0.03) in patient snapshots without a 30-day emergent psychiatric presentation than those with (mean 0.13, SD 0.12) emergent psychiatric presentations.

## DISCUSSION

In this retrospective cohort study of patients receiving outpatient psychiatric care, a machine learning model derived from statewide HIE data showed promising ability to predict 30-day psychiatric ED presentation in a temporally held-out test set. At a prespecified decision threshold, the model maintained 29% precision while identifying over 40% of subsequent psychiatric ED presentations. These findings suggest that structured longitudinal data aggregated across care settings could support short-term psychiatric risk stratification in outpatient practice, with potential to inform proactive outreach and care coordination for patients at elevated near-term risk of decompensation. The limited transportability of the present model should not be interpreted as limiting the broader relevance of the approach. Rather, because psychiatric emergency risk is shaped by local care patterns and data capture, the principal implication of this study is that practices which participate in statewide HIEs may be able to develop locally calibrated models that are more useful than generic risk scores for practice-specific risk stratification.

Previous work has shown that HIE use is independently associated with reduced emergency department length of stay and lower 30-day readmission rates, likely by bridging the information fragmentation that disproportionately affects mental health care.^19,20^ Our findings add to a growing literature on machine learning enabled prediction in mental health care, much of which has focused on suicide-related outcomes^21–23^, treatment response^24^, psychiatric comorbidities^25^, and presence of substance use disorders^26,27^. Several studies have begun to apply machine learning to psychiatric emergencies, attempting to predict ED revisits for psychiatric needs^28^ and detecting aggression among mood disorder patients^29^. In contrast, few studies have examined near-term psychiatric ED presentation in an outpatient psychiatry population, and fewer still have shown potential for utility at a single practice level. The present results suggest that HIE-derived data may offer advantages over single-system records for this task by capturing encounters, diagnoses, and medication information across otherwise fragmented care settings. This may be particularly relevant in psychiatric care, where patients often receive services across multiple institutions and where incomplete visibility into recent utilization patterns can preclude timely intervention.

This study has several salient limitations. First, although the modeling and analytical approach described here may be adaptable to other HIE-enabled settings, the specific model developed here was derived from a single practice population and may not generalize without recalibration or retraining. Second, the outcome relied on structured encounter-type fields and primary diagnosis coding, which may have misclassified some psychiatric emergencies or missed events occurring outside the HIE. Restricting the outcome to primary psychiatric or substance-related diagnoses may also have undercounted crises documented under nonpsychiatric primary codes. Third, because patients could contribute multiple snapshots, performance estimates at the snapshot level may not directly translate to patient-level alerting strategies in clinical deployment. Fourth, subgroup analyses were limited by small event counts in several strata and by missing race and ethnicity data, so fairness-related conclusions remain preliminary. Finally, this study did not evaluate whether acting on model predictions changes patient outcomes, reduces ED use, or is cost-effective in a real-world deployment setting.

## CONCLUSION

In this single-practice retrospective cohort study, a locally developed machine learning model using statewide HIE data predicted 30-day psychiatric ED presentation more accurately than selected general-purpose risk scores. Although the specific model is tailored to one practice and should not be assumed to generalize to other settings, the findings suggest that HIE infrastructure may enable participant practices to develop locally calibrated psychiatric risk models matched to their own patient populations and care environments. Prospective studies are needed to determine whether use of such local models improves care processes or outcomes.

## Supporting information

TripodAIChecklist

## Author contributions

Study concept and design: JLH, ERA; Data acquisition and statistical analyses JLH, ERA; Interpretation of data: All authors; Drafting of manuscript: JLH, ERA; Critical revision of manuscript for important intellectual content: All authors. Supervision: ERA.

## Obtained funding

None

## Data availability statement

Data are available through Big Sky Care Connect with permission. Dr. Havlik takes responsibility for the integrity of the data and the accuracy of the analysis.

## SUPPLEMENTAL MATERIAL

### Methods Supplement

#### Snapshot exclusions

To ensure adequate historical information for feature construction, we required at least 180 days between the patient’s anchor date and the snapshot index date; snapshots not meeting this criterion were excluded. For training snapshots, feature derivation was additionally restricted to data recorded before the prespecified temporal cutoff. Moreover, to approximate a prospective deployment of our model, no snapshots from the training set occurring on or after the June 5, 2023, temporal split were included. This preserved strict temporal separation between the training and test cohorts and reduced the risk of time-correlated leakage.

#### Feature engineering

Utilization features captured prior health care use across 30-, 90-, and 365-day lookback windows, including counts of inpatient, emergency, and outpatient encounters; psychiatric-specific emergency and inpatient encounters; recency of use by setting; recent-use indicators; inpatient length of stay; utilization density; and recent escalation in service use. We also quantified care fragmentation over the prior year using continuity-of-care indices based on the distribution of encounters across providers and facilities.

Diagnostic features were based on ICD-10 codes recorded before the index date and included indicators, counts, and recency measures for any mental disorder diagnosis and for selected psychiatric subgroups, including depressive, anxiety, psychotic, and substance use disorders. Additional diagnosis-derived features captured major nonpsychiatric comorbidity clusters, including cardiovascular, pulmonary, metabolic, renal, hepatic, neurologic, infectious, musculoskeletal, and oncologic conditions.

Pharmacy features were derived from data available in BSCC including continuity of care document medication lists, First Databank data, and Medicaid claims data. Features were classified by rule-based mapping into clinically relevant classes, including antipsychotics, mood stabilizers, antidepressants, benzodiazepines, stimulants, opioids, and medications for addiction treatment. We generated class-specific indicators of recent exposure, recency of fills, and no-fill indicators over 180 days. For antipsychotics and mood stabilizers, we additionally calculated 180-day medication possession ratio and maximum gap in coverage, with derived indicators for suboptimal adherence. Additional medication features captured psychotropic polypharmacy, unique antipsychotic exposure, benzodiazepine-opioid overlap, prescriber fragmentation, and as-needed vs scheduled medication use.

Physiology and laboratory features included the most recent blood pressure and body mass index values and their recency. Laboratory features included recency of any testing, testing intensity relative to encounter volume, and the most recent available values for thyroid function, hemoglobin A1c, and lipid measures. We also derived urine drug screen positivity indicators across multiple lookback windows and recency of the last positive result when identifiable from structured result fields.

Demographic features included age, sex, race, and ethnicity. To incorporate contextual social risk, patient ZIP code prefixes were linked to Social Vulnerability Index and Area Deprivation Index reference data to derive neighborhood-level vulnerability and deprivation measures. We also identified individual-level social risk factors from ICD-10 Z-codes, including homelessness, unemployment, and problems related to primary support group. Finally, because patients could contribute multiple eligible snapshots, we included the number of prior snapshots in the preceding 180 days as a measure of observation intensity.

#### Primary outcome codes

We defined an encounter for psychiatric reasons as an encounter with a primary International Classification of Diseases, Tenth Revision (ICD-10) diagnosis codes for mental, behavioral, or neurodevelopmental disorders (F01-F99), including substance use disorders (F10-F19), and selected symptom or self-harm codes judged *a priori* to represent psychiatric emergency presentations (suicidal ideation [R45.851] and intentional self-harm codes X71-X83).

#### Nested cross-validation for hyperparameter selection

We tuned hyperparameters on and subsequently tested these classifiers’ performance on our training data in a systematic manner using a nested 5 × 5 stratified cross-validation (NCV) framework with a dependent variable of 30-day emergency psychiatric presentation, as defined above. Because patients could contribute multiple snapshots, we used patient-level grouped nested cross-validation to ensure that no patient appeared in both training and validation folds. All categorical variables were one-hot encoded. All predictors were median-imputed and standardized using a robust scaling within a preprocessing pipeline; the pipeline was refit independently inside every resampling split to prevent information leakage. Our inner five-fold cross-validation was used to tune hyperparameters and repeated a grid-search algorithm^30^ and retained the best hyperparameter configuration for that fold.^31^

The inner five-fold cross-validation with grid search for hyperparameter tuning was repeated for five outer folds for each of the three classifier types tested. We conducted preliminary model testing across each of these five outer folds using hyperparameters determined from the inner five-fold cross-validation algorithm. Mean and standard deviation values across the five tuned outer folds provided an estimate of performance for each algorithm at that algorithm’s best-performing hyperparameters. We repeated this testing across our three classifiers. Of the three classifier algorithms undergoing preliminary testing, the classifier with the highest mean outer-fold precision at 33.33% recall at optimized hyperparameter settings proceeded to final model evaluation on unseen test data. We performed an identical operation on our parsimonious clinical baseline model.

#### Trailing 180-day prevalence threshold adjustment

To adjust for recent prevalence of ED visits in the practice, we estimated the most recent 180-day training prevalence immediately prior to the temporal cutoff (π*), and aligned the calibrated decision threshold via a logit shift: threshold applied = expit(logit(threshold from cross-validation) + logit(π*) − logit(π_train)).

#### Final model testing

Once our final model was selected, we tested our hyperparameter-tuned classifier on our temporally-separated testing data, bootstrapping our data 1,000 times with replacement at the patient level to obtain confidence intervals. At the prespecified decision threshold, we recorded the number needed to evaluate and other commonly used classifier metrics of interest including number needed to evaluate, accuracy, sensitivity/recall, specificity, positive predictive value/precision, and negative predictive value. We also reported the overall receiver operator characteristic curve area under curve (ROC AUC) and precision recall curve area under curve (PR AUC) across all decision thresholds, model Brier score, and calibration slope/intercept. To ensure our results were not simply the result of a few patients dominating in snapshot prevalence, we conducted an additional sensitivity analysis using one random snapshot per test patient, and compared metrics qualitatively.

#### Decision curve analysis

Using isotonic calibrated, prior corrected predicted risks (the probabilities used for test decisions), we computed net benefit for each threshold as the proportion of true positives minus the proportion of false positives weighted by the harm:benefit ratio implied by the threshold (w = pt/(1−pt)); treat none (NB = 0) and treat all (NB = prevalence − (1−prevalence)·w) served as reference strategies. We plotted this net benefit as a function of the decision threshold applied, reporting net benefit at the harm:benefit ratio implied by our prespecified decision threshold.

---

**Supplemental Figure 2:**
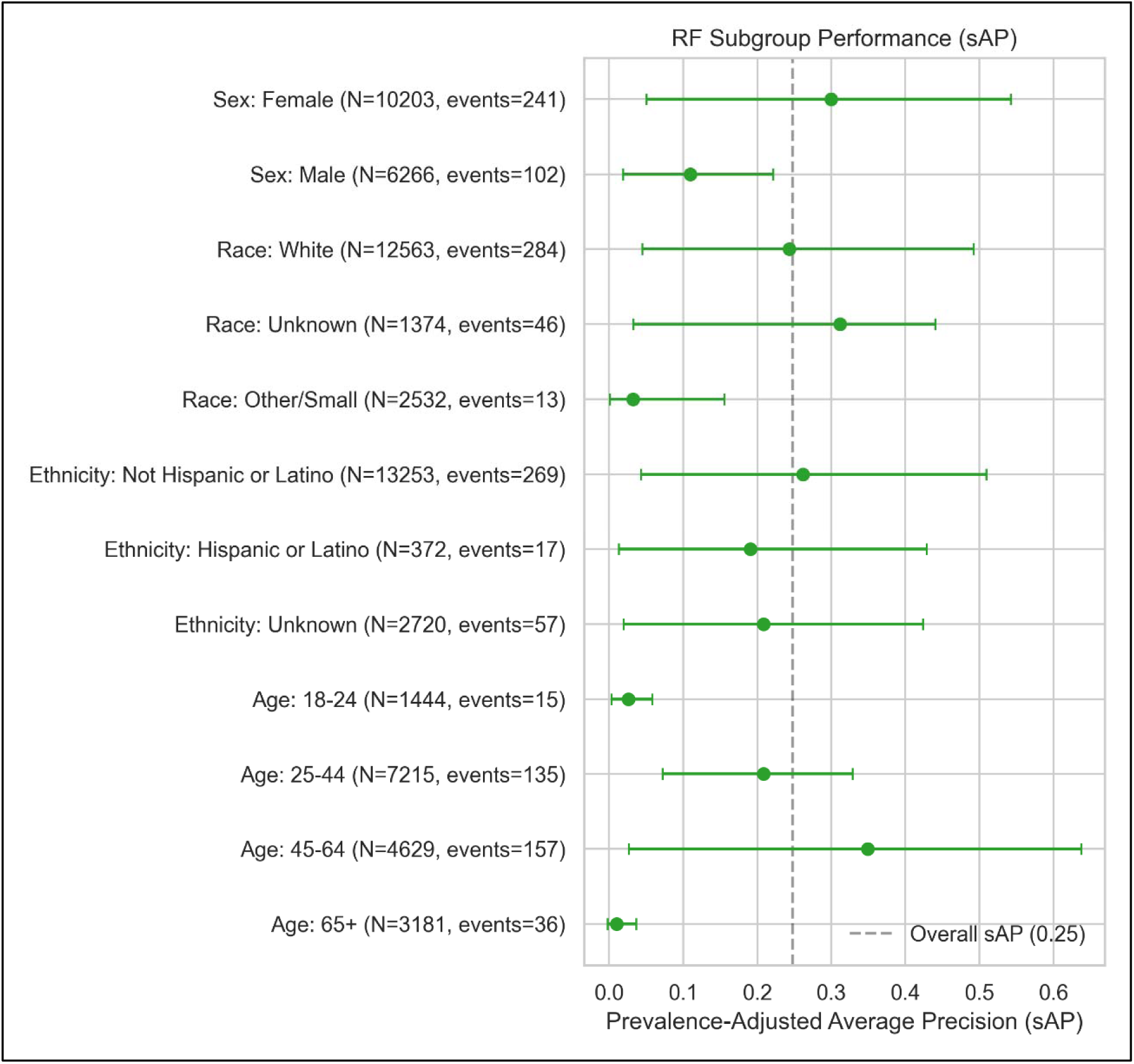
Prevalence-adjusted subgroup analysis, model performance. Prevalence-adjusted analysis for age, sex, and race/ethnicity groups; event denotes a snapshot with a 30-day emergency psychiatric visit. Subgroup performance shown using prevalence adjusted average precision (sAP = [AP − prevalence]/[1 − prevalence]). Points show subgroup sAP with 95% patient-clustered bootstrap confidence intervals. Dashed line denotes overall sAP. Higher values indicate better performance; sAP=0 corresponds to random ranking.

**Supplemental Table 2:** Cross-validated model performance at optimized hyperparameters.

| Fivefold-validated model performance comparison. |  |  |  |  |
| --- | --- | --- | --- | --- |
| **Metric** | **LR** | **RFC** | **LightGBM** | **Parsimonious Clinical Baseline** |
| **Precision at 1/3 Recall** | 0.210 | 0.220 | 0.209 | 0.165 |
| **SD** | 0.067 | 0.049 | 0.048 | 0.039 |
| **ROC AUC** | 0.770 | 0.784 | 0.778 | 0.701 |
| **SD** | 0.037 | 0.024 | 0.024 | 0.018 |
| **PR AUC** | 0.214 | 0.222 | 0.192 | 0.166 |
| **SD** | 0.079 | 0.069 | 0.050 | 0.042 |
| **PPV/Precision** | 0.087 | 0.097 | 0.089 | 0.080 |
| **SD** | 0.018 | 0.022 | 0.012 | 0.014 |
| **Sensitivity/Recall** | 0.660 | 0.644 | 0.663 | 0.524 |
| **SD** | 0.107 | 0.045 | 0.051 | 0.061 |
| **F1 Score** | 0.152 | 0.167 | 0.158 | 0.137 |
| **SD** | 0.026 | 0.033 | 0.018 | 0.019 |
| **Specificity** | 0.760 | 0.794 | 0.777 | 0.796 |
| **SD** | 0.096 | 0.060 | 0.048 | 0.061 |
| **NPV** | 0.986 | 0.986 | 0.986 | 0.981 |
| **SD** | 0.003 | 0.001 | 0.001 | 0.001 |
| **Balanced Accuracy** | 0.709 | 0.719 | 0.720 | 0.660 |
| **SD** | 0.023 | 0.020 | 0.016 | 0.012 |
| ROC: receiver operator characteristic; AUC: area under curve; SD: standard deviation from five outer folds; LR: logistic regression; RFC: random forest classifier; BRFC: balanced random forest classifier; LightGBM: light gradient boosted machine classifier. Highlight denotes top performing model for given metric. |  |  |  |  |

**Supplemental Table 3:** Final Model Metrics.

| Final model performance on temporally separated unseen test data at *a priori* decision threshold. |  |
| --- | --- |
| **Metric** | **Model Performance** |
| **Precision at 1/3 Recall** | 0.294 |
| **SD** | 0.087 |
| **ROC AUC** | 0.850 |
| **SD** | 0.042 |
| **PR AUC** | 0.278 |
| **SD** | 0.111 |
| **Sensitivity/Recall** | 0.492 |
| **SD** | 0.132 |
| **F1 Score** | 0.369 |
| **SD** | 0.104 |
| **Specificity** | 0.975 |
| **SD** | 0.004 |
| **NPV** | 0.989 |
| **SD** | 0.002 |
| **Balanced Accuracy** | 0.734 |
| **SD** | 0.065 |
| ROC: receiver operator characteristic; AUC: area under curve; SD: standard deviation, 1,000x bootstrapped with replacement. |  |

**Supplemental Table 4:** Sensitivity analysis: one random snapshot per patient.

| Final model performance on temporally separated unseen test data at *a priori* decision threshold; one random snapshot per patient. |  |
| --- | --- |
| **Metric** | **Model Performance** |
| **Precision at 1/3 Recall** | 0.192 |
| **SD** | 0.079 |
| **ROC AUC** | 0.919 |
| **SD** | 0.059 |
| **PR AUC** | 0.133 |
| **SD** | 0.076 |
| **Sensitivity/Recall** | 0.333 |
| **SD** | 0.123 |
| **F1 Score** | 0.244 |
| **SD** | 0.088 |
| **Specificity** | 0.988 |
| **SD** | 0.003 |
| **NPV** | 0.994 |
| **SD** | 0.002 |
| **Balanced Accuracy** | 0.661 |
| **SD** | 0.061 |
| ROC: receiver operator characteristic; AUC: area under curve; SD: standard deviation, 1,000x bootstrapped with replacement. |  |

