## Supplemental Table 1 for "A Local Outpatient Practice-Level Prediction Model for Short-Term Psychiatric Emergency Presentation"

**Supplemental Table 1:** Variables included in a 30-day psychiatric emergency prediction model.

| **Description** | **Variable type** |
| --- | --- |
| **Demographics** | |
| Age in years at index date | Continuous |
| Sex | Categorical |
| Race | Categorical |
| Ethnicity | Categorical |
| Age | Continuous |
| Sex | Categorical |
| **Social Determinants of Health** | |
| Any documented homelessness | Binary indicator |
| Days since most recent documented homelessness | Continuous |
| Documented homelessness in prior 90 days | Binary indicator |
| Documented homelessness in prior 365 days | Binary indicator |
| Any documented unemployment | Binary indicator |
| Days since most recent documented unemployment | Continuous |
| Documented unemployment in prior 90 days | Binary indicator |
| Documented unemployment in prior 365 days | Binary indicator |
| Any documented primary support group problems | Binary indicator |
| Days since most recent documented primary support group problems | Continuous |
| Documented primary support group problems in prior 90 days | Binary indicator |
| Documented primary support group problems in prior 365 days | Binary indicator |
| Social Vulnerability Index, housing and transportation theme score | Continuous |
| Social Vulnerability Index, socioeconomic theme score | Continuous |
| Social Vulnerability Index, overall score | Continuous |
| Area Deprivation Index, national percentile rank | Continuous |
| Area Deprivation Index, deprivation score | Continuous |
| Social Vulnerability Index quartile | Categorical |
| **Vital Signs and Anthropometric Measures** | |
| Most recent systolic blood pressure | Continuous |
| Most recent diastolic blood pressure | Continuous |
| Days since most recent blood pressure measurement | Continuous |
| Blood pressure measured in prior 90 days | Binary indicator |
| Blood pressure measured in prior 365 days | Binary indicator |
| Most recent pulse pressure | Continuous |
| Most recent body mass index | Continuous |
| Days since most recent body mass index measurement | Continuous |
| Body mass index measured in prior 90 days | Binary indicator |
| Body mass index measured in prior 365 days | Binary indicator |
| **Healthcare Utilization** | |
| Days since most recent inpatient admission | Continuous |
| Days since most recent emergency department visit | Continuous |
| Days since most recent outpatient visit | Continuous |
| Inpatient admission in prior 90 days | Binary indicator |
| Inpatient admission in prior 365 days | Binary indicator |
| Emergency department visit in prior 90 days | Binary indicator |
| Emergency department visit in prior 365 days | Binary indicator |
| Outpatient visit in prior 90 days | Binary indicator |
| Outpatient visit in prior 365 days | Binary indicator |
| Count of inpatient admissions in prior 30 days | Continuous |
| Count of emergency department visits in prior 30 days | Continuous |
| Count of outpatient visits in prior 30 days | Continuous |
| Count of psychiatric emergency department visits in prior 30 days | Continuous |
| Count of psychiatric inpatient admissions in prior 30 days | Continuous |
| Count of inpatient admissions in prior 90 days | Continuous |
| Count of emergency department visits in prior 90 days | Continuous |
| Count of outpatient visits in prior 90 days | Continuous |
| Count of psychiatric emergency department visits in prior 90 days | Continuous |
| Count of psychiatric inpatient admissions in prior 90 days | Continuous |
| Count of inpatient admissions in prior 365 days | Continuous |
| Count of emergency department visits in prior 365 days | Continuous |
| Count of outpatient visits in prior 365 days | Continuous |
| Count of psychiatric emergency department visits in prior 365 days | Continuous |
| Count of psychiatric inpatient admissions in prior 365 days | Continuous |
| Average inpatient length of stay in prior 365 days | Continuous |
| Maximum inpatient length of stay in prior 365 days | Continuous |
| Inpatient admission rate per year (prior 365 days) | Continuous |
| Inpatient admission rate per year (all available history) | Continuous |
| Days since most recent psychiatric emergency department visit | Continuous |
| Days since most recent psychiatric inpatient admission | Continuous |
| Psychiatric emergency department visit in prior 90 days | Binary indicator |
| Psychiatric emergency department visit in prior 365 days | Binary indicator |
| Psychiatric inpatient admission in prior 90 days | Binary indicator |
| Psychiatric inpatient admission in prior 365 days | Binary indicator |
| Encounter acceleration (30-day rate relative to 365-day rate) | Continuous |
| Encounters per month (prior 365 days) | Continuous |
| Visits per month (prior 365 days) | Continuous |
| Inpatient admissions per month (prior 365 days) | Continuous |
| Emergency department visits per month (prior 365 days) | Continuous |
| Outpatient visits per month (prior 365 days) | Continuous |
| Days since first recorded encounter | Continuous |
| Ratio of inpatient utilization in prior 90 days to preceding 275 days | Continuous |
| Inpatient utilization escalation flag (prior 90 days) | Binary indicator |
| Facility continuity score (prior 365 days) | Continuous |
| **Diagnoses** | |
| Any mental health diagnosis in prior 30 days | Binary indicator |
| Any mental health diagnosis in prior 90 days | Binary indicator |
| Any mental health diagnosis in prior 365 days | Binary indicator |
| Days since most recent mental health diagnosis | Continuous |
| Mental health diagnosis recorded in prior 90 days (count) | Continuous |
| Mental health diagnosis recorded in prior 365 days (count) | Continuous |
| Any depression diagnosis in prior 30 days | Binary indicator |
| Any depression diagnosis in prior 90 days | Binary indicator |
| Any depression diagnosis in prior 365 days | Binary indicator |
| Days since most recent depression diagnosis | Continuous |
| Depression diagnosis recorded in prior 90 days (count) | Continuous |
| Depression diagnosis recorded in prior 365 days (count) | Continuous |
| Any anxiety diagnosis in prior 30 days | Binary indicator |
| Any anxiety diagnosis in prior 90 days | Binary indicator |
| Any anxiety diagnosis in prior 365 days | Binary indicator |
| Days since most recent anxiety diagnosis | Continuous |
| Anxiety diagnosis recorded in prior 90 days (count) | Continuous |
| Anxiety diagnosis recorded in prior 365 days (count) | Continuous |
| Any psychosis diagnosis in prior 30 days | Binary indicator |
| Any psychosis diagnosis in prior 90 days | Binary indicator |
| Any psychosis diagnosis in prior 365 days | Binary indicator |
| Days since most recent psychosis diagnosis | Continuous |
| Psychosis diagnosis recorded in prior 90 days (count) | Continuous |
| Psychosis diagnosis recorded in prior 365 days (count) | Continuous |
| Any substance use disorder diagnosis in prior 30 days | Binary indicator |
| Any substance use disorder diagnosis in prior 90 days | Binary indicator |
| Any substance use disorder diagnosis in prior 365 days | Binary indicator |
| Days since most recent substance use disorder diagnosis | Continuous |
| Substance use disorder diagnosis recorded in prior 90 days (count) | Continuous |
| Substance use disorder diagnosis recorded in prior 365 days (count) | Continuous |
| Any alcohol use disorder diagnosis in prior 30 days | Binary indicator |
| Any alcohol use disorder diagnosis in prior 90 days | Binary indicator |
| Any alcohol use disorder diagnosis in prior 365 days | Binary indicator |
| Days since most recent alcohol use disorder diagnosis | Continuous |
| Alcohol use disorder diagnosis recorded in prior 90 days (count) | Continuous |
| Alcohol use disorder diagnosis recorded in prior 365 days (count) | Continuous |
| Any opioid use disorder diagnosis in prior 30 days | Binary indicator |
| Any opioid use disorder diagnosis in prior 90 days | Binary indicator |
| Any opioid use disorder diagnosis in prior 365 days | Binary indicator |
| Days since most recent opioid use disorder diagnosis | Continuous |
| Opioid use disorder diagnosis recorded in prior 90 days (count) | Continuous |
| Opioid use disorder diagnosis recorded in prior 365 days (count) | Continuous |
| Any hypertension diagnosis in prior 30 days | Binary indicator |
| Any hypertension diagnosis in prior 90 days | Binary indicator |
| Any hypertension diagnosis in prior 365 days | Binary indicator |
| Days since most recent hypertension diagnosis | Continuous |
| Hypertension diagnosis recorded in prior 90 days (count) | Continuous |
| Hypertension diagnosis recorded in prior 365 days (count) | Continuous |
| Any heart failure diagnosis in prior 30 days | Binary indicator |
| Any heart failure diagnosis in prior 90 days | Binary indicator |
| Any heart failure diagnosis in prior 365 days | Binary indicator |
| Days since most recent heart failure diagnosis | Continuous |
| Heart failure diagnosis recorded in prior 90 days (count) | Continuous |
| Heart failure diagnosis recorded in prior 365 days (count) | Continuous |
| Any coronary artery disease diagnosis in prior 30 days | Binary indicator |
| Any coronary artery disease diagnosis in prior 90 days | Binary indicator |
| Any coronary artery disease diagnosis in prior 365 days | Binary indicator |
| Days since most recent coronary artery disease diagnosis | Continuous |
| Coronary artery disease diagnosis recorded in prior 90 days (count) | Continuous |
| Coronary artery disease diagnosis recorded in prior 365 days (count) | Continuous |
| Any atrial fibrillation diagnosis in prior 30 days | Binary indicator |
| Any atrial fibrillation diagnosis in prior 90 days | Binary indicator |
| Any atrial fibrillation diagnosis in prior 365 days | Binary indicator |
| Days since most recent atrial fibrillation diagnosis | Continuous |
| Atrial fibrillation diagnosis recorded in prior 90 days (count) | Continuous |
| Atrial fibrillation diagnosis recorded in prior 365 days (count) | Continuous |
| Any stroke or transient ischemic attack diagnosis in prior 30 days | Binary indicator |
| Any stroke or transient ischemic attack diagnosis in prior 90 days | Binary indicator |
| Any stroke or transient ischemic attack diagnosis in prior 365 days | Binary indicator |
| Days since most recent stroke or transient ischemic attack diagnosis | Continuous |
| Stroke or transient ischemic attack diagnosis recorded in prior 90 days (count) | Continuous |
| Stroke or transient ischemic attack diagnosis recorded in prior 365 days (count) | Continuous |
| Any chronic obstructive pulmonary disease diagnosis in prior 30 days | Binary indicator |
| Any chronic obstructive pulmonary disease diagnosis in prior 90 days | Binary indicator |
| Any chronic obstructive pulmonary disease diagnosis in prior 365 days | Binary indicator |
| Days since most recent chronic obstructive pulmonary disease diagnosis | Continuous |
| Chronic obstructive pulmonary disease diagnosis recorded in prior 90 days (count) | Continuous |
| Chronic obstructive pulmonary disease diagnosis recorded in prior 365 days (count) | Continuous |
| Any asthma diagnosis in prior 30 days | Binary indicator |
| Any asthma diagnosis in prior 90 days | Binary indicator |
| Any asthma diagnosis in prior 365 days | Binary indicator |
| Days since most recent asthma diagnosis | Continuous |
| Asthma diagnosis recorded in prior 90 days (count) | Continuous |
| Asthma diagnosis recorded in prior 365 days (count) | Continuous |
| Any diabetes mellitus diagnosis in prior 30 days | Binary indicator |
| Any diabetes mellitus diagnosis in prior 90 days | Binary indicator |
| Any diabetes mellitus diagnosis in prior 365 days | Binary indicator |
| Days since most recent diabetes mellitus diagnosis | Continuous |
| Diabetes mellitus diagnosis recorded in prior 90 days (count) | Continuous |
| Diabetes mellitus diagnosis recorded in prior 365 days (count) | Continuous |
| Any obesity diagnosis in prior 30 days | Binary indicator |
| Any obesity diagnosis in prior 90 days | Binary indicator |
| Any obesity diagnosis in prior 365 days | Binary indicator |
| Days since most recent obesity diagnosis | Continuous |
| Obesity diagnosis recorded in prior 90 days (count) | Continuous |
| Obesity diagnosis recorded in prior 365 days (count) | Continuous |
| Any hyperlipidemia diagnosis in prior 30 days | Binary indicator |
| Any hyperlipidemia diagnosis in prior 90 days | Binary indicator |
| Any hyperlipidemia diagnosis in prior 365 days | Binary indicator |
| Days since most recent hyperlipidemia diagnosis | Continuous |
| Hyperlipidemia diagnosis recorded in prior 90 days (count) | Continuous |
| Hyperlipidemia diagnosis recorded in prior 365 days (count) | Continuous |
| Any chronic kidney disease diagnosis in prior 30 days | Binary indicator |
| Any chronic kidney disease diagnosis in prior 90 days | Binary indicator |
| Any chronic kidney disease diagnosis in prior 365 days | Binary indicator |
| Days since most recent chronic kidney disease diagnosis | Continuous |
| Chronic kidney disease diagnosis recorded in prior 90 days (count) | Continuous |
| Chronic kidney disease diagnosis recorded in prior 365 days (count) | Continuous |
| Any liver disease diagnosis in prior 30 days | Binary indicator |
| Any liver disease diagnosis in prior 90 days | Binary indicator |
| Any liver disease diagnosis in prior 365 days | Binary indicator |
| Days since most recent liver disease diagnosis | Continuous |
| Liver disease diagnosis recorded in prior 90 days (count) | Continuous |
| Liver disease diagnosis recorded in prior 365 days (count) | Continuous |
| Any epilepsy diagnosis in prior 30 days | Binary indicator |
| Any epilepsy diagnosis in prior 90 days | Binary indicator |
| Any epilepsy diagnosis in prior 365 days | Binary indicator |
| Days since most recent epilepsy diagnosis | Continuous |
| Epilepsy diagnosis recorded in prior 90 days (count) | Continuous |
| Epilepsy diagnosis recorded in prior 365 days (count) | Continuous |
| Any Parkinson disease diagnosis in prior 30 days | Binary indicator |
| Any Parkinson disease diagnosis in prior 90 days | Binary indicator |
| Any Parkinson disease diagnosis in prior 365 days | Binary indicator |
| Days since most recent Parkinson disease diagnosis | Continuous |
| Parkinson disease diagnosis recorded in prior 90 days (count) | Continuous |
| Parkinson disease diagnosis recorded in prior 365 days (count) | Continuous |
| Any dementia diagnosis in prior 30 days | Binary indicator |
| Any dementia diagnosis in prior 90 days | Binary indicator |
| Any dementia diagnosis in prior 365 days | Binary indicator |
| Days since most recent dementia diagnosis | Continuous |
| Dementia diagnosis recorded in prior 90 days (count) | Continuous |
| Dementia diagnosis recorded in prior 365 days (count) | Continuous |
| Any HIV infection diagnosis in prior 30 days | Binary indicator |
| Any HIV infection diagnosis in prior 90 days | Binary indicator |
| Any HIV infection diagnosis in prior 365 days | Binary indicator |
| Days since most recent HIV infection diagnosis | Continuous |
| HIV infection diagnosis recorded in prior 90 days (count) | Continuous |
| HIV infection diagnosis recorded in prior 365 days (count) | Continuous |
| Any hepatitis C diagnosis in prior 30 days | Binary indicator |
| Any hepatitis C diagnosis in prior 90 days | Binary indicator |
| Any hepatitis C diagnosis in prior 365 days | Binary indicator |
| Days since most recent hepatitis C diagnosis | Continuous |
| Hepatitis C diagnosis recorded in prior 90 days (count) | Continuous |
| Hepatitis C diagnosis recorded in prior 365 days (count) | Continuous |
| Any sepsis diagnosis in prior 30 days | Binary indicator |
| Any sepsis diagnosis in prior 90 days | Binary indicator |
| Any sepsis diagnosis in prior 365 days | Binary indicator |
| Days since most recent sepsis diagnosis | Continuous |
| Sepsis diagnosis recorded in prior 90 days (count) | Continuous |
| Sepsis diagnosis recorded in prior 365 days (count) | Continuous |
| Any osteoarthritis diagnosis in prior 30 days | Binary indicator |
| Any osteoarthritis diagnosis in prior 90 days | Binary indicator |
| Any osteoarthritis diagnosis in prior 365 days | Binary indicator |
| Days since most recent osteoarthritis diagnosis | Continuous |
| Osteoarthritis diagnosis recorded in prior 90 days (count) | Continuous |
| Osteoarthritis diagnosis recorded in prior 365 days (count) | Continuous |
| Any rheumatoid arthritis diagnosis in prior 30 days | Binary indicator |
| Any rheumatoid arthritis diagnosis in prior 90 days | Binary indicator |
| Any rheumatoid arthritis diagnosis in prior 365 days | Binary indicator |
| Days since most recent rheumatoid arthritis diagnosis | Continuous |
| Rheumatoid arthritis diagnosis recorded in prior 90 days (count) | Continuous |
| Rheumatoid arthritis diagnosis recorded in prior 365 days (count) | Continuous |
| Any cancer diagnosis in prior 30 days | Binary indicator |
| Any cancer diagnosis in prior 90 days | Binary indicator |
| Any cancer diagnosis in prior 365 days | Binary indicator |
| Days since most recent cancer diagnosis | Continuous |
| Cancer diagnosis recorded in prior 90 days (count) | Continuous |
| Cancer diagnosis recorded in prior 365 days (count) | Continuous |
| **Medications (Pharmacy Fills)** | |
| Active antipsychotic prescription | Binary indicator |
| Antipsychotic fill in prior 90 days | Binary indicator |
| Antipsychotic fill in prior 365 days | Binary indicator |
| No antipsychotic fill in prior 180 days | Binary indicator |
| Medication possession ratio for antipsychotic (prior 180 days) | Continuous |
| Maximum gap in days in antipsychotic therapy (prior 180 days) | Continuous |
| Any gap greater than 30 days in antipsychotic therapy | Binary indicator |
| Medication possession ratio below 0.8 for antipsychotic | Binary indicator |
| Active mood stabilizer prescription | Binary indicator |
| Mood stabilizer fill in prior 90 days | Binary indicator |
| Mood stabilizer fill in prior 365 days | Binary indicator |
| No mood stabilizer fill in prior 180 days | Binary indicator |
| Medication possession ratio for mood stabilizer (prior 180 days) | Continuous |
| Maximum gap in days in mood stabilizer therapy (prior 180 days) | Continuous |
| Any gap greater than 30 days in mood stabilizer therapy | Binary indicator |
| Medication possession ratio below 0.8 for mood stabilizer | Binary indicator |
| Active antidepressant prescription | Binary indicator |
| Antidepressant fill in prior 90 days | Binary indicator |
| Antidepressant fill in prior 365 days | Binary indicator |
| No antidepressant fill in prior 180 days | Binary indicator |
| Active benzodiazepine prescription | Binary indicator |
| Benzodiazepine fill in prior 90 days | Binary indicator |
| Benzodiazepine fill in prior 365 days | Binary indicator |
| No benzodiazepine fill in prior 180 days | Binary indicator |
| Active stimulant prescription | Binary indicator |
| Stimulant fill in prior 90 days | Binary indicator |
| Stimulant fill in prior 365 days | Binary indicator |
| No stimulant fill in prior 180 days | Binary indicator |
| Active opioid prescription | Binary indicator |
| Opioid fill in prior 90 days | Binary indicator |
| Opioid fill in prior 365 days | Binary indicator |
| No opioid fill in prior 180 days | Binary indicator |
| Active medication for addiction treatment prescription | Binary indicator |
| Medication for addiction treatment fill in prior 90 days | Binary indicator |
| Medication for addiction treatment fill in prior 365 days | Binary indicator |
| No medication for addiction treatment fill in prior 180 days | Binary indicator |
| Active psychiatric polypharmacy in prior 90 days | Binary indicator |
| Count of unique antipsychotics in prior year | Count |
| Concurrent benzodiazepine and opioid use in prior 365 days | Binary indicator |
| Average medication possession ratio across drug classes | Continuous |
| Distinct medication classes per month (prior 365 days) | Continuous |
| Count of active psychiatric medications (prior 30 days) | Count |
| Count of active psychiatric medications (prior 90 days) | Count |
| Days of concurrent psychiatric medication use (prior 90 days) | Continuous |
| Count of unique prescribers (prior 365 days) | Count |
| Prescriber continuity score (prior 365 days) | Continuous |
| Any active as-needed (PRN) medication | Binary indicator |
| Ratio of as-needed (PRN) to scheduled medications | Continuous |
| **Laboratory Values** | |
| Days since most recent laboratory test of any type | Continuous |
| Any laboratory test in prior 90 days | Binary indicator |
| Any laboratory test in prior 365 days | Binary indicator |
| Laboratory tests per encounter (prior 365 days) | Continuous |
| Most recent thyroid stimulating hormone value | Continuous |
| Days since most recent thyroid stimulating hormone measurement | Continuous |
| Thyroid stimulating hormone measured in prior 90 days | Binary indicator |
| Thyroid stimulating hormone measured in prior 365 days | Binary indicator |
| Most recent hemoglobin A1c value | Continuous |
| Days since most recent hemoglobin A1c measurement | Continuous |
| Hemoglobin A1c measured in prior 90 days | Binary indicator |
| Hemoglobin A1c measured in prior 365 days | Binary indicator |
| Most recent free thyroxine (T4) value | Continuous |
| Days since most recent free thyroxine (T4) measurement | Continuous |
| Free thyroxine (T4) measured in prior 90 days | Binary indicator |
| Free thyroxine (T4) measured in prior 365 days | Binary indicator |
| Most recent free triiodothyronine (T3) value | Continuous |
| Days since most recent free triiodothyronine (T3) measurement | Continuous |
| Free triiodothyronine (T3) measured in prior 90 days | Binary indicator |
| Free triiodothyronine (T3) measured in prior 365 days | Binary indicator |
| Most recent HDL cholesterol value | Continuous |
| Days since most recent HDL cholesterol measurement | Continuous |
| Most recent triglycerides value | Continuous |
| Days since most recent triglycerides measurement | Continuous |
| Most recent LDL cholesterol value | Continuous |
| Days since most recent LDL cholesterol measurement | Continuous |
| HDL cholesterol measured in prior 90 days | Binary indicator |
| HDL cholesterol measured in prior 365 days | Binary indicator |
| Triglycerides measured in prior 90 days | Binary indicator |
| Triglycerides measured in prior 365 days | Binary indicator |
| LDL cholesterol measured in prior 90 days | Binary indicator |
| LDL cholesterol measured in prior 365 days | Binary indicator |
| Total cholesterol measured in prior 90 days | Binary indicator |
| Total cholesterol measured in prior 365 days | Binary indicator |
| Most recent total cholesterol to HDL ratio value | Continuous |
| Days since most recent total cholesterol to HDL ratio measurement | Continuous |
| Total cholesterol to HDL ratio measured in prior 90 days | Binary indicator |
| Total cholesterol to HDL ratio measured in prior 365 days | Binary indicator |
| Triglyceride to HDL ratio (most recent) | Continuous |
| **Urine Drug Screen** | |
| Any positive urine drug screen in prior 30 days | Binary indicator |
| Any positive urine drug screen in prior 90 days | Binary indicator |
| Any positive urine drug screen in prior 365 days | Binary indicator |
| Positive urine drug screen in prior 90 days (recency indicator) | Binary indicator |
| Positive urine drug screen in prior 365 days (recency indicator) | Binary indicator |
| ADI: Area Deprivation Index; BMI: body mass index; CAD: coronary artery disease; COPD: chronic obstructive pulmonary disease; ED: emergency department; HDL: high-density lipoprotein; HIV: human immunodeficiency virus; LDL: low-density lipoprotein; PRN: pro re nata (as needed); SVI: Social Vulnerability Index; T3: triiodothyronine; T4: thyroxine. Continuous features were median-imputed and standardized with a robust scaler; categorical features were one-hot encoded. All time windows are referenced to the prediction index date. | |
